# Estimating data-driven COVID-19 mitigation strategies for safe university reopening

**DOI:** 10.1101/2021.08.13.21261983

**Authors:** Qihui Yang, Don M. Gruenbacher, Caterina M. Scoglio

## Abstract

After one pandemic year of remote or hybrid instructional modes, universities in the United States are now planning for an in-person fall semester in 2021. However, it is uncertain what the vaccination rate will look like after students, faculty, and staff return to campus. To help inform university-reopening policies, we collected survey data on social contact patterns and developed an agent-based model to simulate the spread of COVID-19 in university settings. In this paper, we aim to identify the immunity threshold that, if exceeded, would lead to a relatively safe on-campus experience for the university population. With relaxed non-pharmaceutical interventions, we estimated that immunity in at least 60% of the university population is needed for safe university reopening. Still, attention needs to be paid to extreme events that could lead to huge infection size spikes. At an immune level of 60%, continuing non-pharmaceutical interventions, such as wearing masks, could lead to an 89% reduction in the maximum cumulative infection, which reflects the possible non-negligible infection size from extreme events.

## 1. Introduction

Since March 2020, most U.S. universities have suspended in-person operations and employed remote or hybrid instructional modes in response to the unprecedented restrictions caused by the coronavirus disease 2019 (COVID-19). Buoyed by the wider vaccine availability, a growing number of universities in the United States have announced full reopening with an in-person fall semester in 2021. Despite the high effectiveness of COVID-19 vaccines, many institutions have not required a vaccine mandate. In addition, studies have suggested that COVID vaccination hesitancy appears to be high in certain population subgroups such as young adults [1–3]. For example, Sharma et al. [3] reported that 47.5% of participants were hesitant to get vaccinated based on questionnaires distributed to college students in a Southern U.S. University. As there will be people returning to campus through out-of-state or international travel, it is hard to project the vaccination rate in the coming fall semester, which is crucial for the epidemic forecast.

Facing such challenges, universities struggle with plans in resuming normal operations while mitigating the risks of severe acute respiratory syndrome coronavirus 2 (SARS-CoV-2). Since the pandemic started, over 700,000 cases have been reported concerning American colleges and universities [4]. Several agent-based models have been developed to guide decision-making on testing frequencies, mask usage, social distancing, and class sizes during school reopening [5–8]. Asgary et al. developed an agent-based simulation tool to evaluate the impact of different testing strategies and protective measures such as masking [5]. Junge et al. [6] analyzed the herd immunity threshold needed for safe university reopening and found that vaccine coverage over 80% makes it possible to resume in-person instructions safely. Due to data unavailability, many of these models simulate contacts based on random mixing or assumptions regarding class schedules and common locations. Previous studies have shown that contact networks could significantly impact the accuracy of epidemic predictions and the effectiveness of control strategies [9–11], highlighting the importance of data collection on real-world contact patterns.

COVID-19 is still evolving, and researchers are devoted to retrieving its epidemiological parameters [12–14]. Recent studies have reported non-exponential distributions for critical transition times between different disease states, such as the infectious period [15,16]. Whereas most epidemic models have been developed based on Markovian processes with transition times following exponential distributions. Such unrealistic assumptions could impair the accuracy of model predictions, and non-Markovian models that accept arbitrary distributions for the transition times of the individual between different compartments have started to draw attention from scholars [17–19].

In this study, we develop an agent-based model to examine the mitigation strategies needed for safe university reopening in the 2021 fall semester. The model incorporates a social contact network based on survey data. Considering a highly effective vaccine, we simulate COVID spreading in a university population under two scenarios: 1) relaxation of non-pharmaceutical interventions (NPIs) and 2) adoption of NPIs, such as wearing masks. The outcomes are valuable to understand the impact of initial immune levels on the future epidemic spread, thereby helping inform university-reopening policies.

The contributions of this paper are summarized as follows:

- We develop an agent-based model incorporating non-Markovian transition times and real contact networks based on survey data.
- We estimate that immunity in at least 60% of the university population is needed to ensure a healthy campus with relaxed NPIs.
- We observe that the implementation of NPIs can dramatically reduce the maximum cumulative infection, and continued NPIs are recommended to mitigate risks from extreme events.

## 2. Survey data

To parametrize the model, we conducted a social contact survey administered to all students, faculty, and staff at Kansas State University between December 2, 2020, and January 25, 2021. We sent emails to 6,196 faculty and staff members, and 20,755 students, and received responses from 3,581 participants with a success rate of 13.29%. The survey data contain information about age segment, role at the university, housing status, and number of close contacts categorized by duration ranges in a week. We also gathered information about visit frequency, duration, and the number of contacts at different locations.

Figure 1 depicts the age, housing status, and the number of close contacts in a week by duration ranges. We can see that 50.70% of participants reported being in the age category 18–24, followed by 15.90% of participants in the age category 25–35. Regarding housing status, 9.82% of students live in sororities or fraternities, 21.00% live in on-campus housing, and the remaining students live in off-campus apartments or houses. The majority of faculty and staff (98.91%) choose off-campus housing options. In the survey, we explicitly indicate that examples of contacts regularly met more than 4 hours per week are roommates, family members, or coworkers. Contacts between 1 to 4 hours per week may refer to friends or classmates, and contacts between 15 minutes to 1 hour per week could be friends or others that the participant might occasionally meet. Overall, the contact patterns categorized by role reveal that students living in sororities or fraternities have more contacts while faculty and staff have fewer contacts. For example, regarding the duration of more than 4 hours per week (yellow), the median number of contacts for students living at sororities or fraternities is 8 contacts compared to faculty and staff with 2 contacts. More statistics about the survey data can be found in the supplementary material. (c) The number of contacts the survey respondent regularly meets categorized by duration ranges in a week and the role of the survey respondent

**Figure 1.**
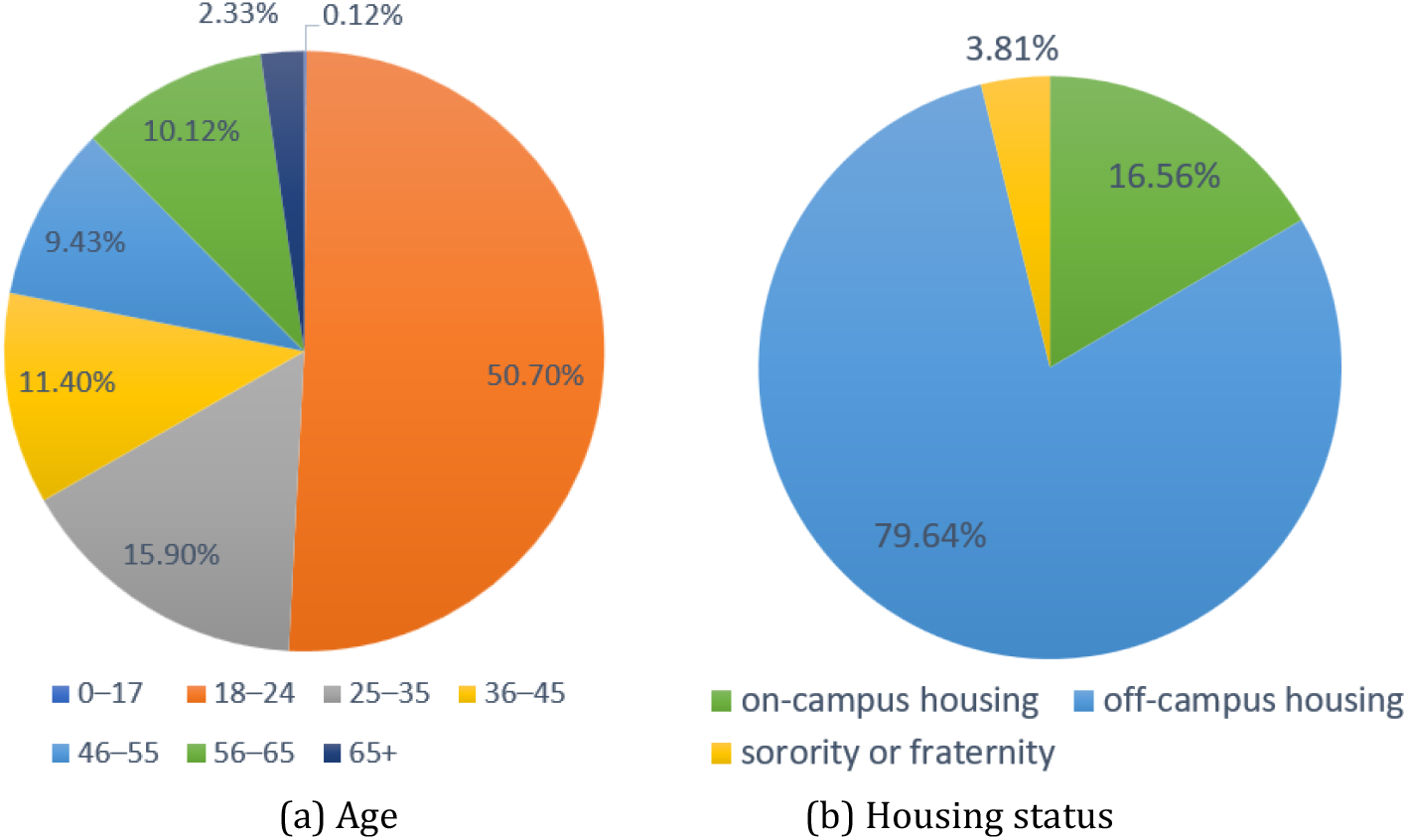
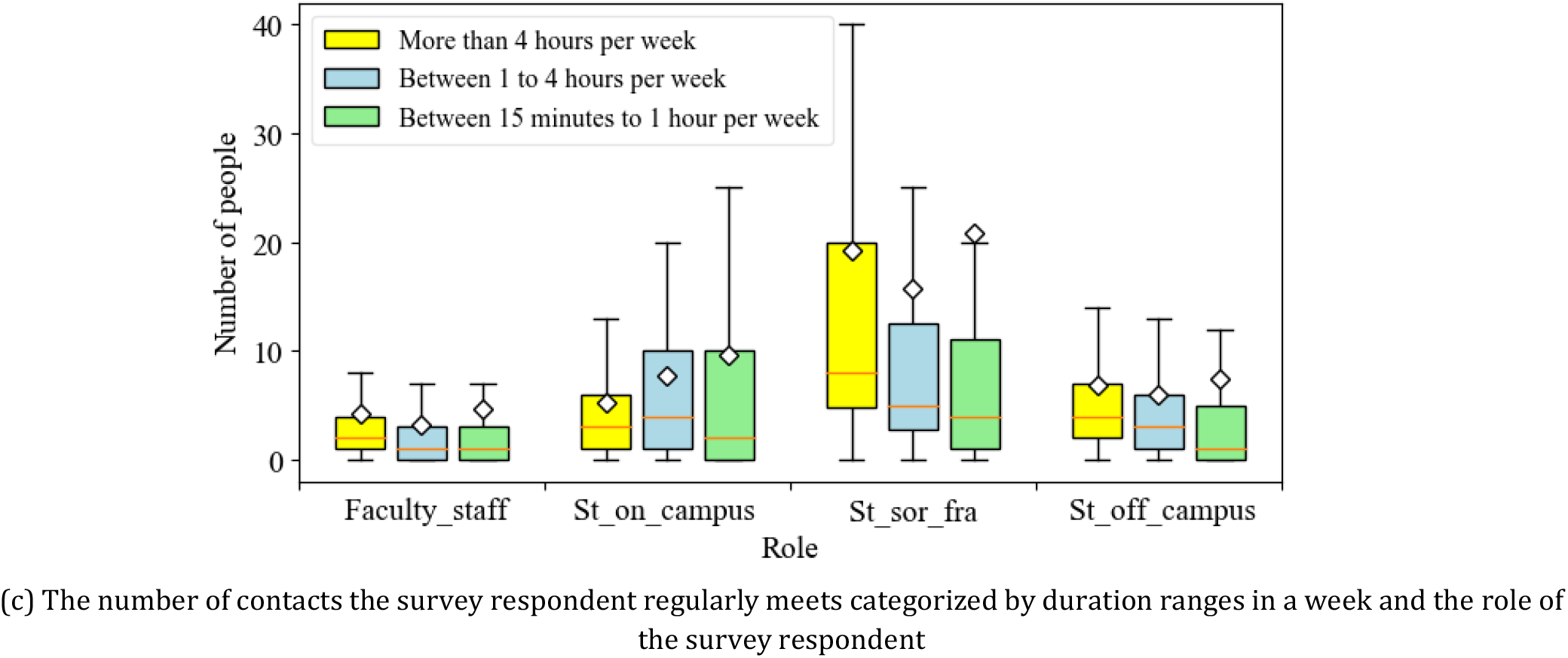
Population characteristics and the number of contacts per person differentiated by duration ranges in a week of the survey respondent. *Faculty_staff, St_on_campus, St_sor_fra, St_off_campus* represent the following four types of roles: faculty and staff; students living on-campus housing; students living in a sorority or fraternity; and students living-off campus.

## 3. Model

The model developed mainly consists of two types of agents, namely person and location agents. During initialization, we create 26,000 person-agents, representing 20,000 students and 6,000 faculty and staff. Each person-agent is assigned to one role category of a) faculty and staff, b) student living on campus, c) student living off campus, or d) student living at a sorority or fraternity. With each role category, the age attribute is associated with each person-agent based on the age distribution obtained from the survey data. Each location-agent plays one role of: a) recreation center or any gym or other shared exercise spaces; b) union, dining centers, and coffee shops on campus; c) bars, restaurants, and coffee shops off campus; d) stores and other types of services off campus; and e) other types of social gathering such as sport, religious, and social events.

### Daily activities

Figure 2 depicts the structure of the person-agent. Each person-agent has contact with people it regularly meets in the three contact lists, and meets other people at the five types of locations. More specifically, the people within each person’s contact lists are randomly selected from the whole population during model initialization, and remain unchanged throughout each simulation run. By contrast, the people each person meets at specific locations are dynamic, as the contact will only occur if two agents are within the same location. At different times of the day, each person agent visits these locations based on the visit frequency, the duration, and the number of contacts, which are sampled from exponential distributions with parameters retrieved from the survey data.

**Figure 2.**
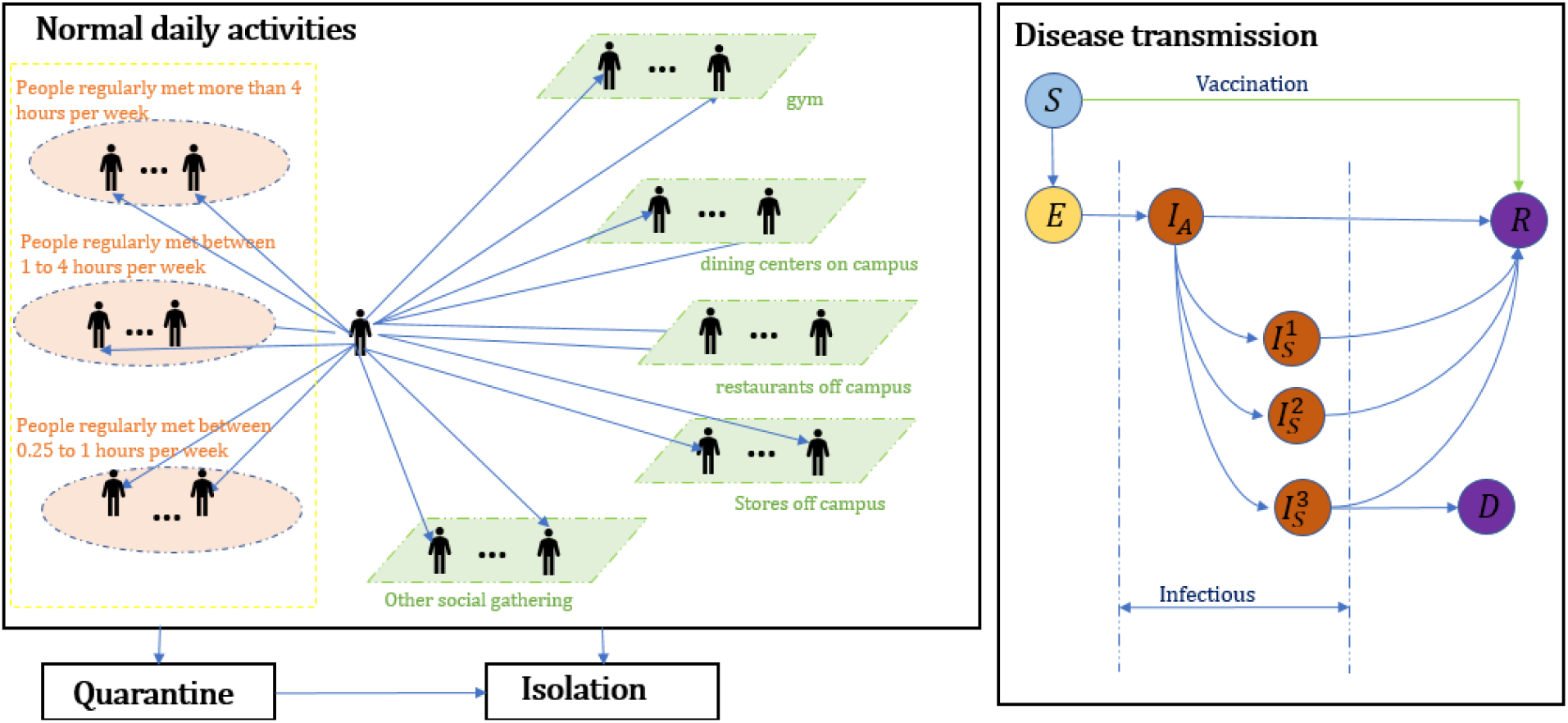
Structure of the person-agent in the model.

### Disease transmission

In line with reference [20,21], each individual has a state reflecting its health status: susceptible (*S*), exposed (*E*, infected but not infectious), asymptomatic (*I*_*A*_), symptomatic differentiated by mild 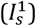, severe 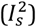 and critical illness 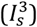, recovered (*R*), and dead (*D*). Contacting an infectious individual, a susceptible individual could become infected and transition to the exposed state based on the transmission probability per contact *β*. An individual in the exposed state will transition to the infectious state *I*_*A*_ after a period lognormal distributed with a mean of 4.6 days and a standard deviation of 4.8 days [22–25]. Infected individuals may develop symptoms based on age-dependent probabilities [26,27]. The probabilities that symptomatic cases develop into mild, severe, or critical illness are also age-dependent [27]. The length of time for an individual to transition from states *I*_*A*_ to symptoms onset (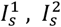 or 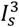) follows a lognormal distribution with a mean of 1 day and standard deviation of 0.9 days [16]. The recovery time for asymptomatic cases and mild symptomatic cases is a period sampled from a lognormal distribution with a mean of 8 days and a standard deviation of 2 days [28]. Individuals with severe and critical illness recover after a period sampled from a lognormal distribution with a mean of 14 days and a standard deviation of 2.4 days [26]. Accounting for outside transmissions due to non-university contacts, we assume the university population has few contacts with local communities, and more details can be found in the supplementary material.

### Testing, quarantine, and isolation

When an individual has mild symptoms (state 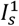), it may get a COVID test based on a 70% probability. Individuals in the critical or moderate symptomatic state (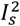 or 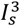) may get tested with a 95% probability [8]. The delay for returning test results is uniformly distributed between 0 and 2 days. An individual will be isolated for ten days after being tested positive. If the individual shows symptoms, the time spent in isolation will count starting from its first day showing symptoms. In addition, we assume that 50% of contacts of a confirmed case can be identified via contact tracing and thus be quarantined. During the quarantine period, the agent will be isolated if the person shows symptoms and is tested positive. The agent will stop daily activities in both quarantine and isolation states.

### Assumptions for estimating the herd immunization threshold

We consider a two-dose vaccination allocated to susceptible people with a time interval of 28 days. The vaccination rollout rate is uniformly distributed between 0 and 10 doses per day. Vaccine efficacy is 92% for the first dose and 95.6% for the second dose [29]. Vaccine efficacy is modelled as the probability that a vaccinated susceptible agent directly transitions to the recovered state. To initialize the epidemics, we randomly select 30 individuals and infect them with the transmission probability *β*. During scenario analyses, the initial immunization level *α* is the percentage of people in the recovered state at the start of the simulation, representing the presumably achieved immunity either through prior infection or vaccination. We perform 500 simulation runs for each scenario, and record the number of active cases and cumulative infected cases in each simulation run.

## 4 Results and discussions

In this section, we present and compare simulation results under scenarios of NPI relaxation (wearing masks and maintaining social distancing are optional) and NPI adoption (wearing masks and maintaining social distancing are mandatory). In each scenario, we vary the initial immunization level, *α*, which refers to the percentage of people having immunity at the start of the fall semester.

### 4.1 NPI relaxation

Considering relaxed NPIs, we assume the COVID-19 transmission probability *β* as 0.03 per contact, following relevant studies [20,30,31]. Figure 3 shows the average percentage of active cases over time with 95 percent confidence intervals given different initial immune levels. While our main focus is to examine the epidemic dynamics over the fall semester (the grey shaded area in figure 3), we depict the simulation results from August 17, 2021 – June 12, 2022 to provide a relatively complete picture of the epidemic curve. For *α* = 40%, the percentage of active cases by the end of the semester is 3.20% (95% CI: 3.08% – 3.32%). In contrast, when more than 60% of the population initially has immunity, disease outbreaks can be controlled relatively well. At the time of writing, vaccination rates in some U.S. states remain to be low. For example, only 36.0% of the state’s population is fully vaccinated in Arkansas. The simulation results show that such a relatively low vaccination rate could lead to a large number of infections, under relaxed NPIs.

**Figure 3.**
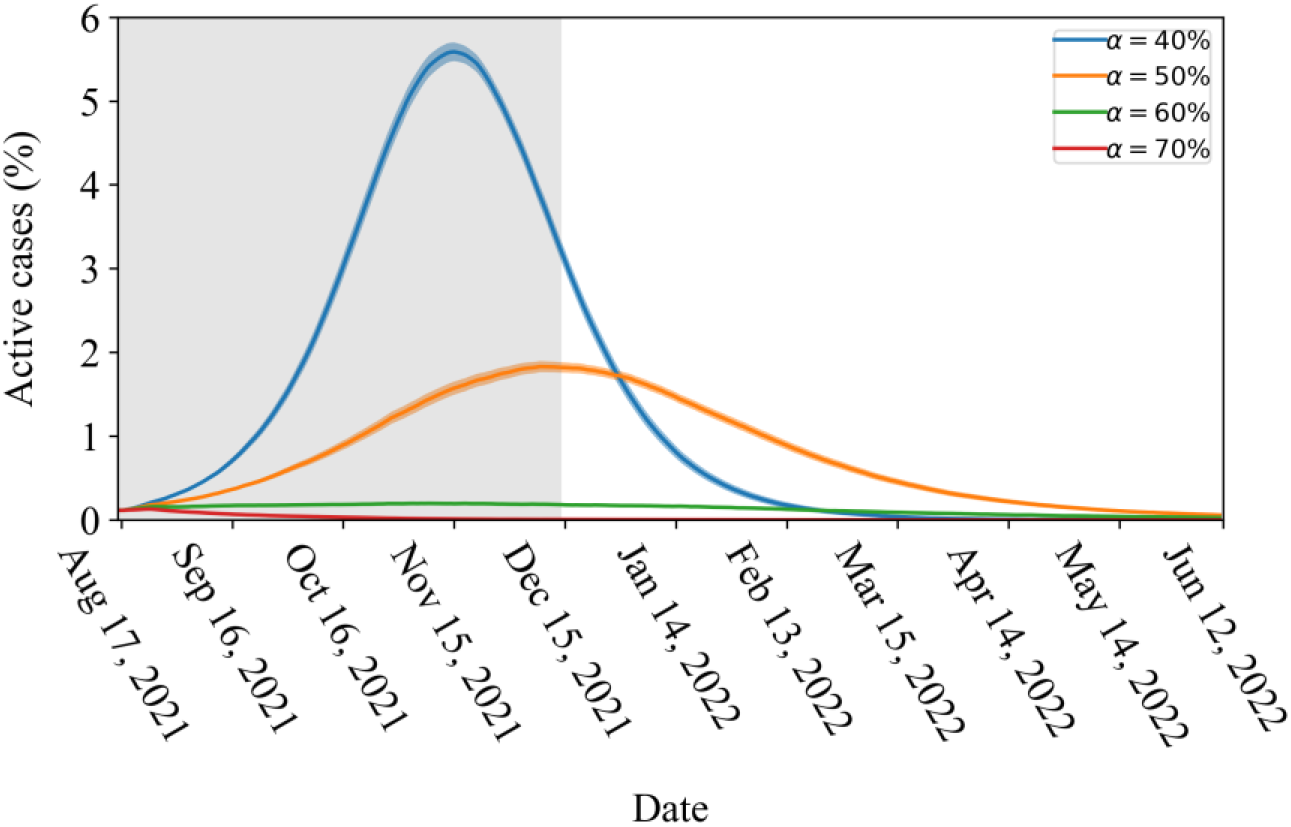
The percentage of active cases for scenarios with initial immunization level *α* varied between 40% and 70%, with an interval of 10%. While our main focus is to examine the epidemic dynamics over the coming fall semester (the grey shaded area), we provide a relatively complete picture of the epidemic curve from August 17, 2021 – June 12, 2022.

Under the same simulation settings as in figure 3, figure 4 shows more plots with *α* varied between 50% and 64% with an interval of 2%. We can see that the epidemic size is quite sensitive to the changes in *α*. The average percentage of active cases by the end of the semester decreases dramatically from 1.82% (95% CI: 1.76% – 1.88%) for *α* = 50% to 0.18% (95% CI: 0.16% – 0.20%) for *α* = 60%.

**Figure 4.**
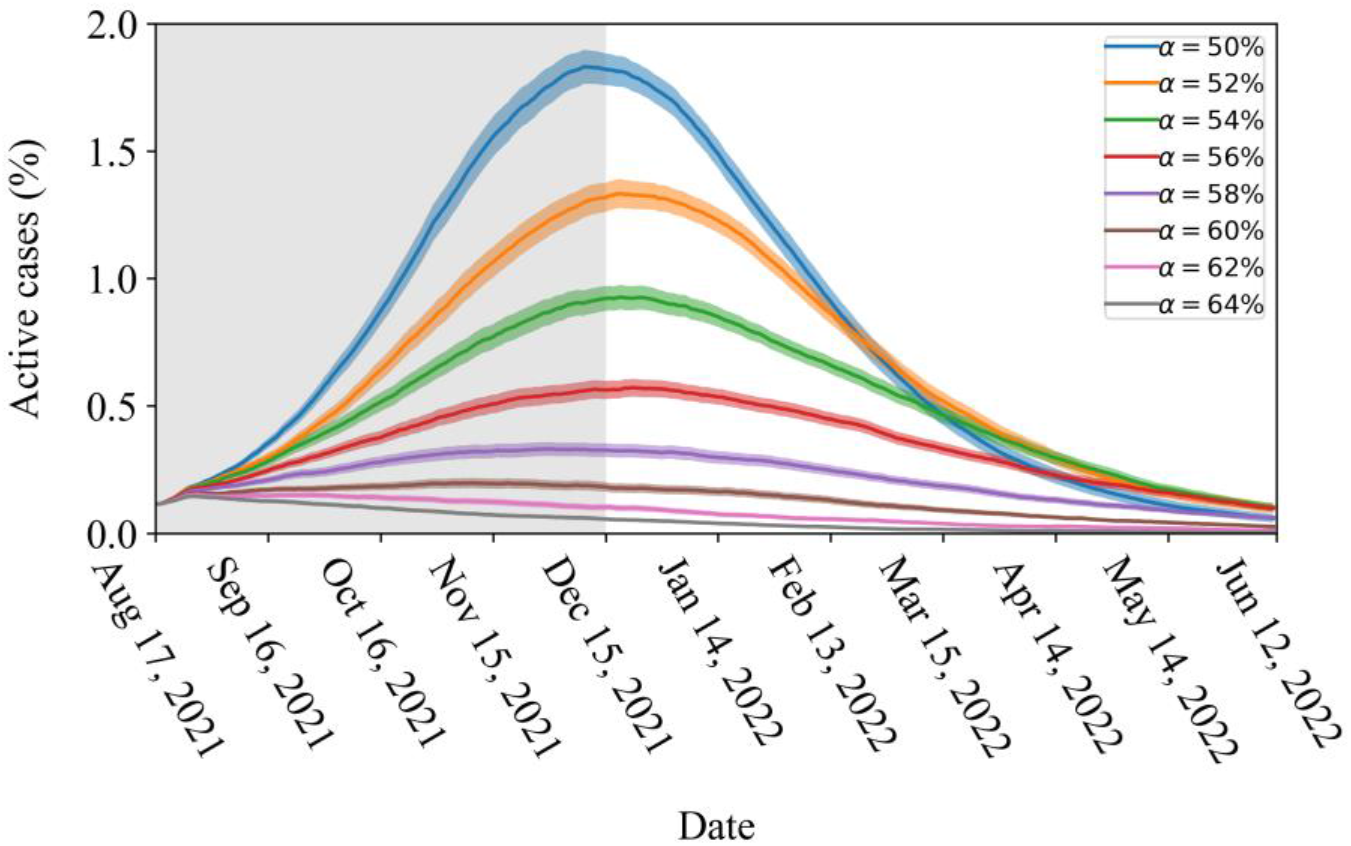
The percentage of active cases for scenarios with initial immunization level *α* varied between 50% and 64% with an interval of 2%. While our main focus is to examine the epidemic dynamics over the coming fall semester (the grey shaded area), we provide a relatively complete picture of the epidemic curve from August 17, 2021 – June 12, 2022.

We also calculate the peak time and height of the peak infection for the median number of active cases, and the results are listed in Table 1. Interestingly, we can see that as *α* increases from 50% to 60%, the peak of infection reduces dramatically from 1.94% to 0.15% of the total population, and slightly declines after *α*=60%. When the initial immune level is above 60%, the peak occurs at day 8 – 10 and the epidemic is well contained.

**Table 1.** The peak infection and the peak time for the median number of active cases. The beginning of the simulation corresponds to the start of the fall semester. Peak time is the number of days elapsed after the initial infection.

| Initial immune level $\alpha$ | Peak value | Peak time (days) |
| --- | --- | --- |
| 40% | 5.90% | 90 |
| 50% | 1.94% | 116 |
| 52% | 1.41% | 125 |
| 54% | 0.93% | 125 |
| 56% | 0.56% | 147 |
| 58% | 0.26% | 122 |
| 60% | 0.15% | 10 |
| 62% | 0.14% | 9 |
| 64% | 0.14% | 9 |
| 70% | 0.13% | 8 |

As additional information, figure 5 displays boxplots for the cumulative infected cases and active cases by the end of the fall semester. Though the median percentages of active cases for *α*=62% and *α*=64% range from 0.02% – 0.05%, there are multiple outliers over 0.50% a day. Similarly, the cumulative infections at *α*=62% and *α*=64% have median values of 1.22% and 0.91%, respectively, but have multiple outliers over 5.00% of the university population. For *α*=70%, at which the median value for the active cases equals 0, the maximum cumulative infected cases can reach up to 3.09% of the population. Though such undesired outcomes rarely happen, it suggests that superspreading events might result in large outbreaks if NPIs are not put in place.

**Figure 5.**
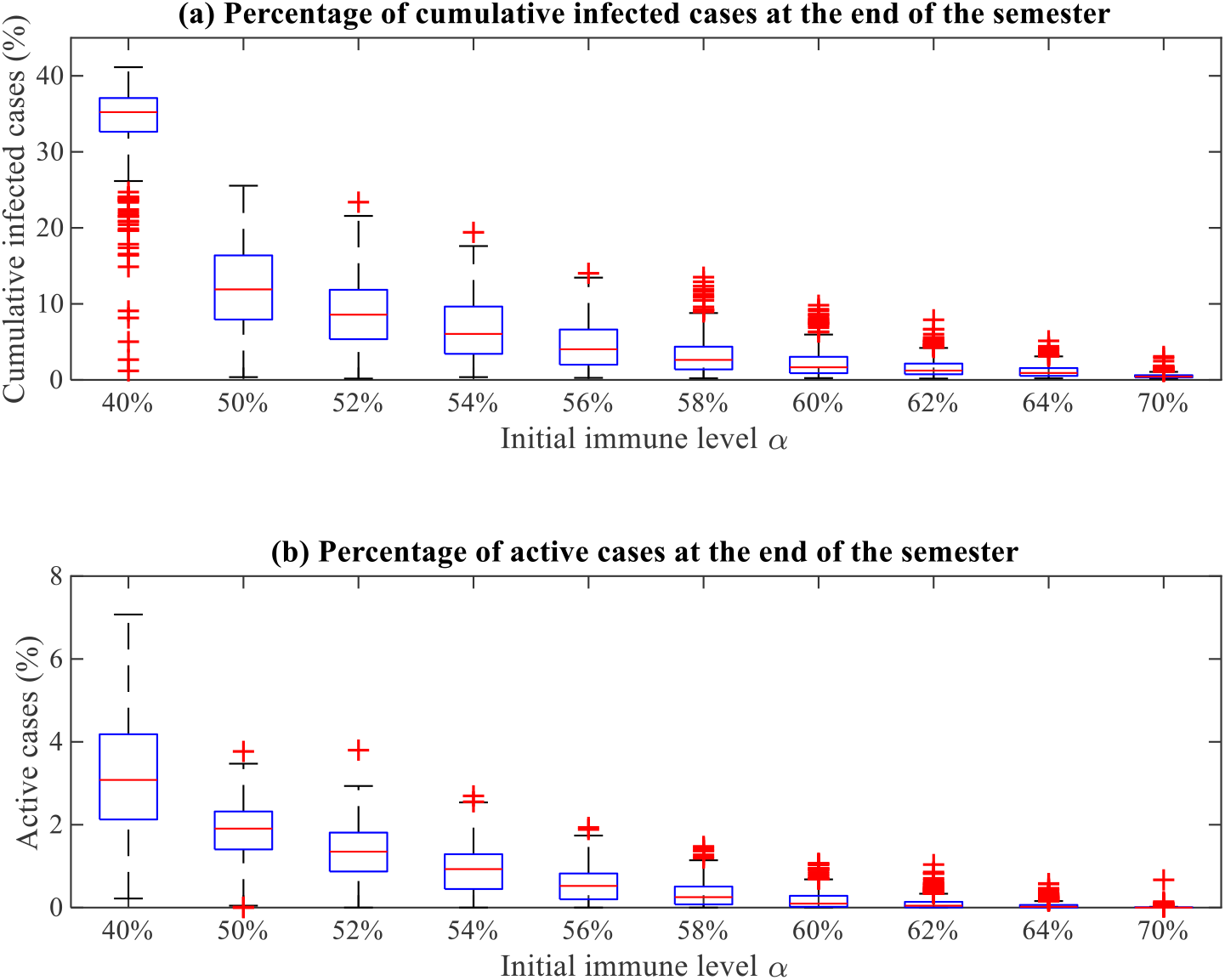
Distributions of the cumulative infected cases and active cases in the percentage of the total population by the end of the fall semester with the relaxation of NPIs.

### 4. 2 NPI adoption

As reported in [32–34], NPIs such as wearing masks could reduce the infection likelihood by at by at least 50%. Accordingly, we set β = 0.015 for scenarios with NPIs. Figure 6 displays the percentage of active cases given different initial immune levels, *α*. Compared to simulation results with relaxed NPIs, we observe that NPIs cause a dramatic reduction in the epidemic size. Over 30% of the population initially immune could ensure relatively safe university reopening, but extra care should still be paid attention to extreme events. If more than 35% of the population initially has immunity, the median percentage of active cases falls below 0.02%. The median percentages of active cases for 50% – 70% all equal 0, while the median percentages of cumulative infected cases range from 0.20% – 0.37%. Compared to scenarios with relaxed NPIs, continuing NPIs can substantially reduce risks from large outbreaks. For instance, the maximum cumulative infected cases reduce from 9.82% (figure 5) to 1.07% (figure 6) for *α* = 60%.

**Figure 6.**
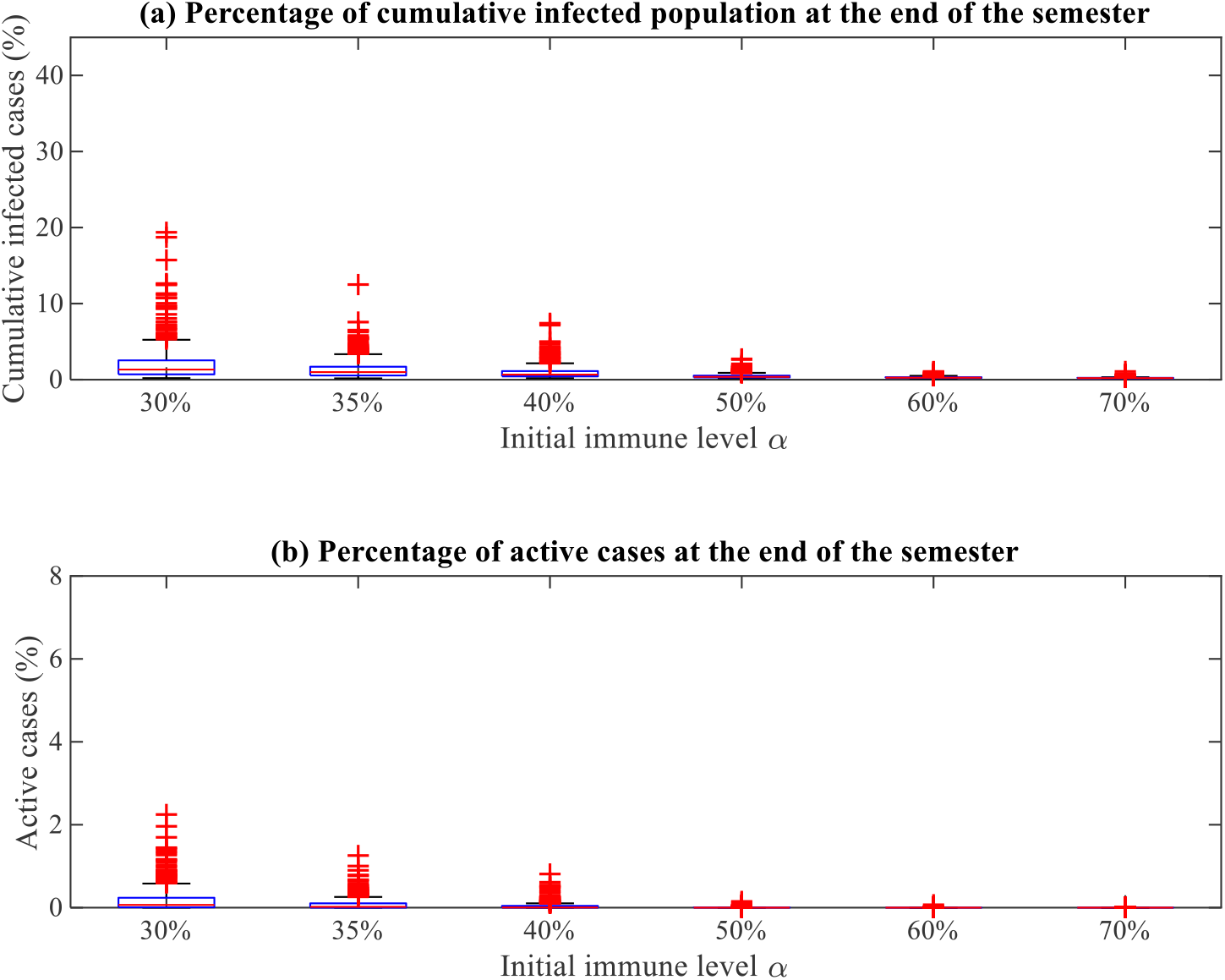
Distributions of cumulative infected cases and active cases in the percentage of the total population by the end of the fall semester with the adoption of NPIs.

## 5. Conclusions

As the fall semester is approaching, many universities have announced the plan for an in-person semester, with relaxed COVID-19 related guidelines. Without a vaccination mandate policy, the exact immune level of the university population is largely unknown, considering that many students, faculty, and staff are coming back through out-of-state or international travel. Our simulation results suggest that, on average, a vaccination rate at 60% of the university population can lead to safe university reopening with social distancing policies lifted. However, it should note that attention still needs to be paid to the superspreading events that may lead to large infections, even with an immune level of 70%. However, an immune level below 60% may pose significant risks to the public health of the university population with the relaxation of NPIs.

Since the COVID outbreak, studies have shown that encouraging people to get vaccinated and continuing non-pharmaceutical control policies are effective ways to suppress the disease spread [35–37]. Consistently, we conclude that it is possible to ensure a healthy campus community associated with NPIs even at a lower immune level of 30%. At the same initial immune level of 60%, reducing the infection rate by half could lead to an 89.10% reduction concerning the maximum cumulative infections, which reflects the possible non-negligible infection from extreme events. Therefore, it is recommended that people continue to exercise social distancing measures for the coming fall semester.

In this study, we developed an agent-based disease transmission model based on a real contact network structure and non-Markovian transition time distributions. We draw our conclusions based on the early estimations of transmission probabilities during the pandemic. Sustaining NPIs will become even more critically important, given the highly contagious delta variant, which could lead to worse scenarios. One limitation of our model lies in the difficulty in setting the parameters due to data unavailability. Therefore, we make several assumptions and conduct more simulations, the results of which can be found in the supplementary material. We find that the number of infections used to initialize the outbreak and vaccination rollout rate has only a minimal impact on the initial immune level needed for a safe reopening. Nonetheless, we believe that the outcome from this study provides important messages for universities planning for a full reopening, especially those located in the states with low vaccination rates.

## Data Availability

All data generated or analyzed during this study are included in this manuscript and supplementary material.

## Ethics statement

The Committee on Research Involving Human Subjects / Institutional Review Board (IRB) for Kansas State University has determined that the research is EXEMPT from further IRB review.

## Authors’ contributions

Q.Y. performed the numerical studies and wrote the first draft of the manuscript; D.M.G. and C.M.S. supervised the research, edited and revised the manuscript; all the authors contributed to the research design and interpretation of the results.

