## supplementary material for "Estimating data-driven COVID-19 mitigation strategies for safe university reopening"

This appendix presents more details about model parameters and simulation results.

### S1. Model parameters

In the tables below, *Faculty_staff*, *St_on_campus*, *St_sor_fra*, *St_off_campus* represent the following four types of agent roles: faculty and staff; students living on-campus housing; students living at a sorority or fraternity; and students living-off campus. Table S1 shows the age distribution for each agent role category.

**Table S1. Proportion of agents by age group and role**

| Agent role | Age category | Proportion of agents distributed according to the survey data |
|---|---|---|
| St_on_campus | Less than 18 | 0.0096 |
| | 18-24 | 0.9873 |
| | 25-35 | 0.0032 |
| St_off_campus | 18-24 | 0.7266 |
| | 25-35 | 0.1928 |
| | 36-45 | 0.0482 |
| | 46-55 | 0.0254 |
| | 56-65 | 0.0061 |
| | 65+ | 0.0009 |
| St_sor_fra | 18–24 | 1.0000 |
| Faculty_staff | 18–24 | 0.0247 |
| | 25–35 | 0.1854 |
| | 36–45 | 0.2444 |
| | 46–55 | 0.2219 |
| | 56–65 | 0.2626 |
| | 65+ | 0.0611 |

Table S2 shows the statistics about the number of contacts a person regularly meets categorized by duration ranges in a week, corresponding to figure 1(c) in the main manuscript.

**Table S2. The number of people the survey respondent regularly meets in a week**

| Total contact durations | Median (IQR) | Mean | Role type |
|---|---|---|---|
| More than 4 hours per week (examples are roommates, family members, or coworkers) | 3 (1–6) | 5.22 | St_on_campus |
| | 4 (2–7) | 6.85 | St_off_campus |
| | 8 (4.5–20) | 19.21 | St_sor_fra |
| | 2 (1–4) | 4.17 | Faculty_staff |
| Between 1 to 4 hours per week (for instance friends or classmates) | 4 (1–10) | 7.73 | St_on_campus |
| | 3 (1–6) | 5.93 | St_off_campus |

| | | | |
|---|---|---|---|
| | 5 (2.5–13) | 15.74 | St_sor_fra |
| | 1 (0–3) | 3.20 | Faculty_staff |
| Between 15 minutes to 1 hour per week (for instance friends or others that the survey respondent occasionally meets) | 2 (0–10) | 9.57 | St_on_campus |
| | 1 (0–5) | 7.38 | St_off_campus |
| | 4 (1–12) | 20.88 | St_sor_fra |
| | 1 (0–3) | 4.69 | Faculty_staff |

IQR: Interquartile range.

Figure S1 depicts boxplots of visit frequency, duration, number of contacts at the five types of locations. Similar across all role type of the participant, fewest contacts occurred at rec or any gym or other shared exercise spaces, followed by union, dining centers and coffee shops on campus. Tables S3–S5 provide median (interquartile range) and mean values corresponding to the boxplots in figure S1.

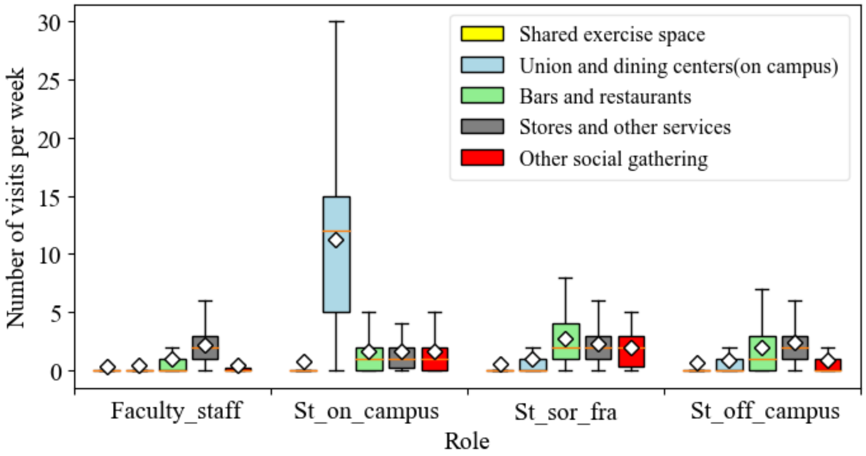


(a) Reported number of visits to each type of location per week of each survey respondent

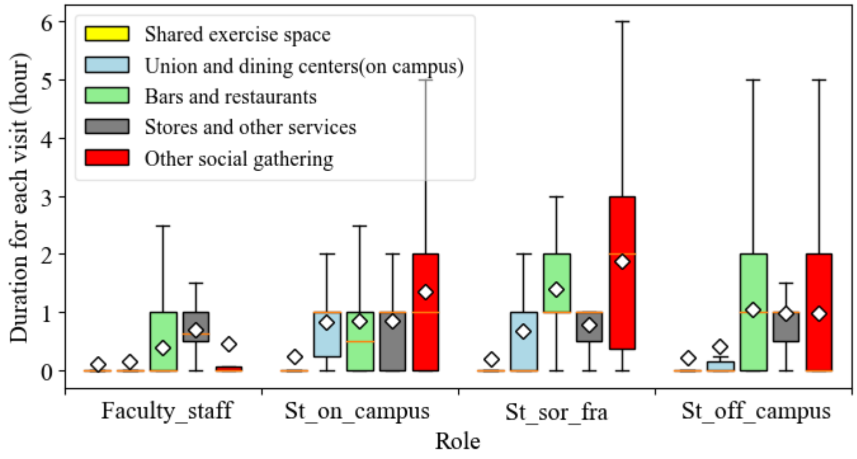


(b) Reported duration for each visit in hours of each survey respondent

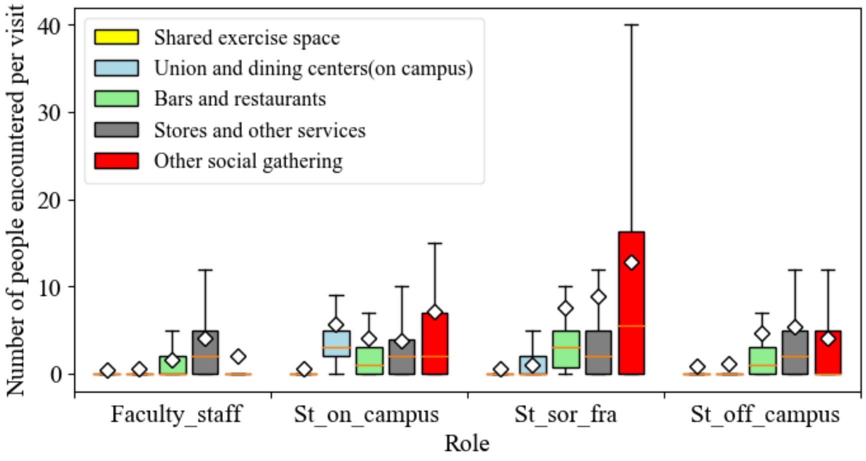


(c) Reported number of people each survey respondent encountered in each visit

**Figure S1. Boxplots of social contact patterns at five types of locations.**

**Table S3. The reported number of visits to each type of location per week of each survey respondent**

| Location | Median (IQR) | Mean | Role type |
|---|---|---|---|
| Rec or any Gym or other shared exercise space | 0 (0–0) | 0.77 | St_on_campus |
| | 0 (0–0) | 0.66 | St_off_campus |
| | 0 (0–0) | 0.55 | St_sor_fra |
| | 0 (0–0) | 0.32 | Faculty_staff |
| Union, dining centers, and coffee shops on campus | 12 (5–15) | 11.31 | St_on_campus |
| | 0 (0–1) | 0.85 | St_off_campus |
| | 0 (0–1) | 1.01 | St_sor_fra |
| | 0 (0–0) | 0.47 | Faculty_staff |
| Bars, restaurants, and coffee shops off campus | 1 (0–2) | 1.68 | St_on_campus |
| | 1 (0–3) | 1.93 | St_off_campus |
| | 2 (1–4) | 2.69 | St_sor_fra |
| | 0 (0–1) | 0.96 | Faculty_staff |
| Stores and other type of services off campus | 1 (0–2) | 1.60 | St_on_campus |
| | 2 (1–3) | 2.42 | St_off_campus |
| | 2 (1–3) | 2.25 | St_sor_fra |
| | 2 (1–3) | 2.22 | Faculty_staff |
| Other types of social gathering (examples are sport, religious, and social events) | 1 (0–2) | 1.63 | St_on_campus |
| | 0 (0–1) | 0.90 | St_off_campus |
| | 2 (0.25–3) | 1.95 | St_sor_fra |
| | 0 (0–0.3) | 0.39 | Faculty_staff |

**Table S4. The reported duration for each visit in hours at different locations**

| Location | Median (IQR) | Mean | Role type |
|---|---|---|---|
| Rec or any Gym or other shared exercise space | 0 (0–0) | 0.23 | St_on_campus |
| | 0 (0–0) | 0.21 | St_off_campus |
| | 0 (0–0) | 0.20 | St_sor_fra |
| | 0 (0–0) | 0.11 | Faculty_staff |
| Union, dining centers, and coffee shops on campus | 1 (0.25–1) | 0.83 | St_on_campus |
| | 0 (0–0.15) | 0.41 | St_off_campus |
| | 0 (0–1) | 0.69 | St_sor_fra |

| | | | |
|---|---|---|---|
| | 0 (0–0) | 0.16 | Faculty_staff |
| Bars, restaurants, and coffee shops off campus | 0.5 (0–1) | 0.86 | St_on_campus |
| | 1 (0–2) | 1.04 | St_off_campus |
| | 1 (1–2) | 1.40 | St_sor_fra |
| | 0 (0–1) | 0.40 | Faculty_staff |
| Stores and other type of services off campus | 1 (0–1) | 0.85 | St_on_campus |
| | 1 (0.5–1) | 0.99 | St_off_campus |
| | 1 (0.5–1) | 0.80 | St_sor_fra |
| | 0.5 (0.45–1) | 0.70 | Faculty_staff |
| Other types of social gathering (examples are sport, religious, and social events) | 1 (0–2) | 1.34 | St_on_campus |
| | 0 (0–2) | 0.99 | St_off_campus |
| | 2 (0.25–3) | 1.88 | St_sor_fra |
| | 0 (0–0.13) | 0.46 | Faculty_staff |

**Table S5. The reported number of contacts each survey respondent encounters in each visit**

| Location | Median (IQR) | Mean | Role type |
|---|---|---|---|
| Rec or any Gym or other shared exercise space | 0 (0–0) | 0.65 | St_on_campus |
| | 0 (0–0) | 0.94 | St_off_campus |
| | 0 (0–0) | 0.60 | St_sor_fra |
| | 0 (0–0) | 0.41 | Faculty_staff |
| Union, dining centers, and coffee shops on campus | 3 (2–5) | 5.71 | St_on_campus |
| | 0 (0–0) | 1.23 | St_off_campus |
| | 0 (0–2) | 1.03 | St_sor_fra |
| | 0 (0–0) | 0.52 | Faculty_staff |
| Bars, restaurants, and coffee shops off campus | 1 (0–3) | 4.03 | St_on_campus |
| | 1 (0–3) | 4.59 | St_off_campus |
| | 3 (0.5–5) | 7.58 | St_sor_fra |
| | 0 (0–2) | 1.56 | Faculty_staff |
| Stores and other type of services off campus | 2 (0–4) | 3.78 | St_on_campus |
| | 2 (0–5) | 5.36 | St_off_campus |
| | 2 (0–5) | 8.95 | St_sor_fra |
| | 2 (0–5) | 4.10 | Faculty_staff |
| Other types of social gathering (examples are sport, religious, and social events) | 2 (0–7) | 7.07 | St_on_campus |
| | 0 (0–5) | 4.07 | St_off_campus |
| | 5.5 (0–17.5) | 12.88 | St_sor_fra |
| | 0 (0–0) | 1.98 | Faculty_staff |

### Table S6. Other model parameters

| | Age group category | | | | | | | | | |
|---|---|---|---|---|---|---|---|---|---|---|
| Description | 0–9 | 10–19 | 20–29 | 30–39 | 40–49 | 50–59 | 60–69 | 70–79 | 80+ | Source |
| Probability of developing symptoms | 0.5 | 0.55 | 0.6 | 0.65 | 0.7 | 0.75 | 0.8 | 0.85 | 0.9 | [1,2] |
| Proportion of symptomatic cases requiring hospitalization | 0.00 | 0.00 | 0.01 | 0.03 | 0.05 | 0.10 | 0.17 | 0.24 | 0.27 | [2] |
| Proportion of hospitalized cases requiring critical care | 0.05 | 0.05 | 0.05 | 0.05 | 0.06 | 0.12 | 0.27 | 0.43 | 0.71 | [2] |
| Infection fatality Ratio (probability of death from a critical case) | 0.00002 | 0.00006 | 0.00030 | 0.00080 | 0.00150 | 0.00600 | 0.02200 | 0.05100 | 0.09300 | [2] |
| Time to transition from Exposed state to infectiousness onset (e.g. viral shedding begins) | Lognormal distribution with mean=4.6 days and std=4.8 days | | | | | | | | | [3–6] |
| Time from infectious onset to showing symptoms | Lognormal distribution with mean=1.0 days and std=0.9 days | | | | | | | | | [7,8] |
| Time to transition from states $I_A$ to $R$ | Lognormal distribution with mean=8.0 days and std=2.0 days | | | | | | | | | [9] |
| Time to transition from states $I_s^1$ to $R$ | Lognormal distribution with mean=8.0 days and std=2.0 days | | | | | | | | | [9] |
| Time to transition from states $I_s^2$ to $R$ | Lognormal distribution with mean=14.0 days and std=2.4 days | | | | | | | | | [1] |
| Time to transition from states $I_s^3$ to $R$ | Lognormal distribution with mean=14.0 days and std=2.4 days | | | | | | | | | [1] |
| False negative rate during testing | 0.13 | | | | | | | | | [10] |
| Delay in receiving test results | Uniform distribution [0, 2] days | | | | | | | | | Assumed |
| Probability for opting to be tested | 0.7 for mild cases (agents in state $I_s^1$), 0.95 for moderate and critical cases (agents in state $I_s^2$ and state $I_s^3$) | | | | | | | | | [11] |
| Percentage of contacts that can be traced | 50% | | | | | | | | | Assumed |

### 2. Simulation results

#### (1) Scenarios under different vaccination rollout rates

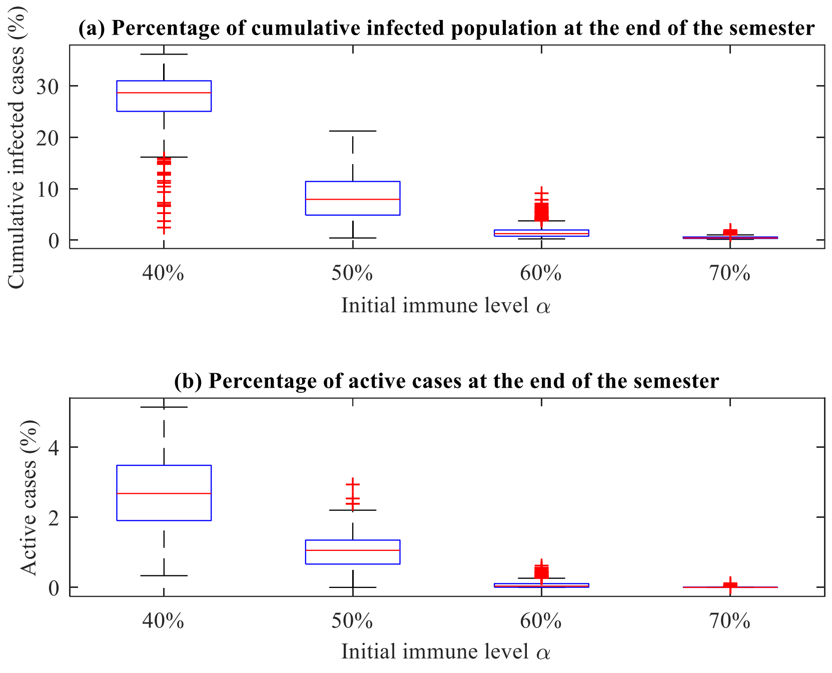


**Figure S2. Simulation results under relaxed non-pharmaceutical interventions with vaccination rate uniformly distributed between** [0, 50]

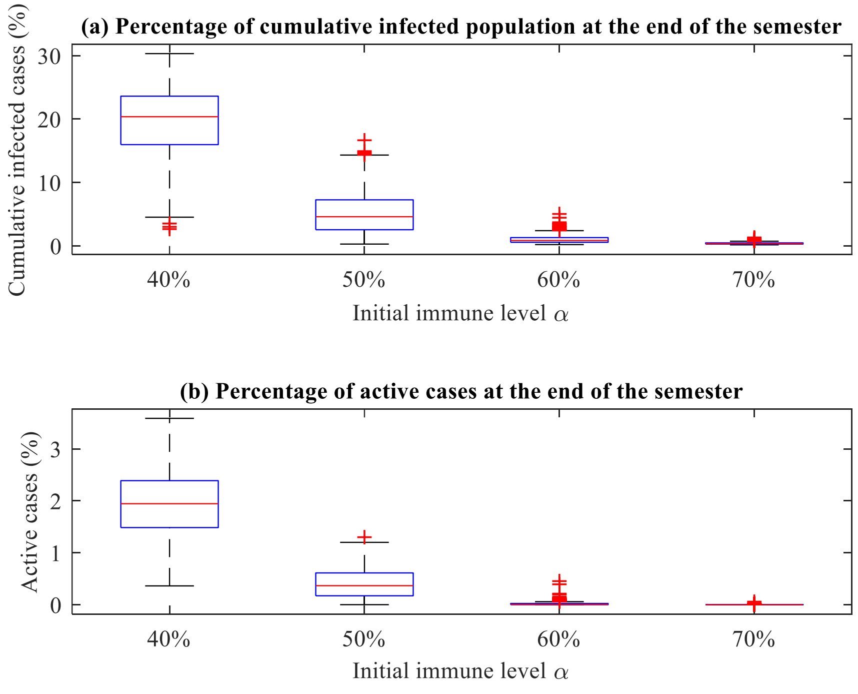


**Figure S3. Simulation results under relaxed non-pharmaceutical interventions with vaccination rate uniformly distributed between** [0, 100]

**(2) Scenarios selecting 15 people to initialize the epidemic**

β = 0.03, rollout rate is uniformly distributed between [0, 10]

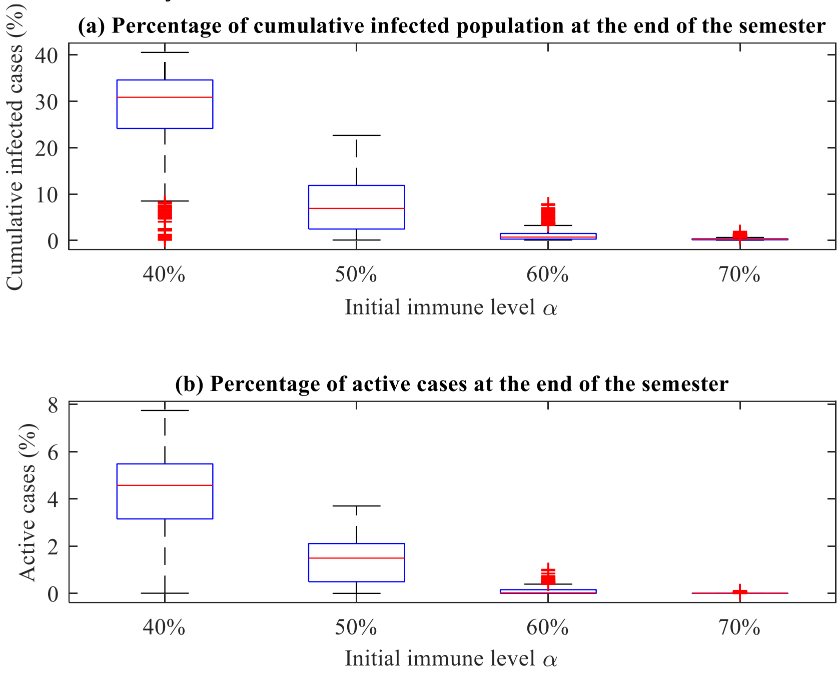


**Figure S4. Simulation results under relaxed non-pharmaceutical interventions with vaccination rate uniformly distributed between** [0, 10]
